# Resistome characterization of extended-spectrum beta-lactamase (ESBL)-producing *Escherichia coli* isolated from wastewater treatment utilities in Oregon

**DOI:** 10.1101/2021.11.15.21266365

**Authors:** Maeghan Easler, Clinton Cheney, Jared D. Johnson, Marjan Khorshidi Zadeh, Jacquelynn N. Nguyen, Sue Yee Yiu, Joy Waite-Cusic, Tyler S. Radniecki, Tala Navab-Daneshmand

## Abstract

Infections resistant to broad spectrum antibiotics due to the emergence of extended-spectrum beta-lactamase (ESBL)-producing Enterobacteriaceae is of global concern. This study characterizes the resistome (i.e., entire ecology of resistance determinants) of 11 ESBL-producing *Escherichia coli* isolates collected from eight wastewater treatment utilities across Oregon. Whole genome sequencing was performed to identify the most abundant antibiotic resistance genes including ESBL-associated genes, virulence factors, as well as their sequence types. Moreover, the phenotypes of antibiotic resistance were characterized. ESBL-associated genes (i.e., *bla*_CMY_, *bla*_CTX_, *bla*_SHV_, *bla*_TEM_) were found in all but one of the isolates with five isolates carrying two of these genes (4 with *bla*_CTX_ and *bla*_TEM_; 1 with *bla*_CMY_ and *bla*_TEM_). The *ampC* gene and virulence factors were present in all the *E. coli* isolates. Across all the isolates, 31 different antibiotic resistance genes were identified. Additionally, all *E. coli* isolates harbored phenotypic resistance to beta-lactams (penicillins and cephalosporins), while eight of the 11 isolates carried multi-drug resistance phenotypes (resistance to three or more classes of antibiotics). Findings highlight the risks associated with the presence of ESBL-producing *E. coli* isolates in wastewater systems that have the potential to enter the environment and may pose direct or indirect risks to human health.

## INTRODUCTION

Emergence of bacterial infections resistance to antibiotics, including beta-lactams, is a global public health threat. Resistance to broad-spectrum beta-lactams, such as ampicillin and 3^rd^ and 4^th^ generation cephalosporins, by extended-spectrum beta-lactamase (ESBL)-producing Enterobacteriaceae is an emerging concern even in areas with high restrictions on antibiotic consumption, such as Norway (Jørgensen *et al*. 2017). Infections with ESBL-producing Enterobacteriaceae are shown to increase the risk of hospitalization, likelihood of discharge to a chronic care facility, and in-hospital mortality (Schwaber *et al*. 2006). ESBL-producing species are characterized by their ability to hydrolyze a broad spectrum of beta-lactam antibiotics, including oxyimino cephalosporins, monobactams, and penicillins. ESBLs are confirmed as the resistance mechanism by verifying that beta-lactamase inhibitors, clavulanic acid or tazobactam, restore the sensitivity to the beta-lactam (Bush and Fisher 2011). Development of novel beta-lactams has been ineffective to reduce the spread of ESBL-producers; in fact, selective pressure of novel antibiotics has resulted in the evolution of more pernicious beta-lactamases (Bradford 2001).

ESBL-associated genes are found in many species of Enterobacteriaceae, especially in *Escherichia coli*, a primary indicator of fecal bacterial contamination (Palucha et al. 1999). Among clinical isolates of ESBL-producing *E. coli* from various countries, including Indonesia, Iran, Korea, and the United States, *bla*_CTX-M_ has been reported as the most common ESBL gene with *bla*_CTX-M-15_ the most prevalent subtype (Park et al. 2009; Sidjabat et al. 2009; Severin et al. 2010; Haghighatpanah et al. 2016). In addition, many of the ESBL-producing *E. coli* carry virulence factors that can enhance their pathogenicity (Jiang *et al*. 2019). An example of a virulence factor is tellurite resistance, encoded by genes such as *terC*, which has been implicated in bacterial resistance to phagocytosis and oxidative stress and contributing to prolonged urinary tract infections (Valková *et al*. 2007; Turkovicova *et al*. 2016). Moreover, ESBL genes are often encoded on plasmids (i.e., mobile genetic elements) and hence, can easily spread via horizontal gene transfer (Haenni *et al*. 2018; Liu *et al*. 2019). In addition, ESBL-producers are commonly resistant to other antibiotics, including gentamicin, sulfamethoxazole-trimethoprim, and ciprofloxacin, resulting in fewer clinical treatment options (Schwaber *et al*. 2005). Furthermore, ESBL-producing *E. coli* are reportedly more likely to be multi-drug resistance (i.e., resistant to three or more classes of antibiotics) than non ESBL-producers (Blaak *et al*. 2015; Jørgensen *et al*. 2017). The resistome (i.e., entire ecology of resistance determinants) of ESBL-producing *E. coli* is not clearly understood and further knowledge is needed to enable development of treatment options for ESBL-producing *E. coli* infections.

Although ESBL-producers have been primarily observed and described in clinical settings, ESBL genes and the bacterial strains that harbor them are also found in many environmental reservoirs including agricultural soils, livestock, and surface water (Ben Said *et al*. 2015; Blaak *et al*. 2015; Haenni *et al*. 2018). Wastewater treatment plants are also major reservoirs and sources of antibiotic-resistant bacteria and their determinant antibiotic resistance genes (ARGs) (Manaia *et al*. 2018; Alexander *et al*. 2020). Recently, ESBL-producing *E. coli* were detected in all the samples collected in a wastewater monitoring study in Germany (Schmiege *et al*. 2021). Similar to clinical settings, *bla*_CTX-M_ genes are common ESBL determinants in wastewater systems (Paulshus et al. 2019; Schages et al. 2020). Most of the studies on ESBL-associated genes in wastewater systems are from Europe; however, there is a limited knowledge regarding ESBL-producing *E. coli* in wastewater utilities in the United States. With growing attention to wastewater-based epidemiology as an important public health surveillance framework, understanding the resistome of ESBL-producing *E. coli* in wastewater treatment systems is critical (Riquelme *et al*. 2021).

This study characterizes the resistome of 11 ESBL-producing *E. coli* isolates collected from various wastewater treatment utilities across the state of Oregon. We used whole genome sequencing (WGS) to determine the antibiotic resistance genotypes of ESBL-producing *E. coli* isolates, their sequence types (STs), ARGs including ESBL-associated genes as well as virulence factors. Moreover, the phenotypes of antibiotic resistance were characterized. The significance and originality of this study is resistome characterization of ESBL-producing *E. coli* isolates in the U.S. wastewater treatment systems that have the potential to enter the environment and pose risks to human health.

## MATERIALS AND METHODS

### *E. coli* isolation and antibiotic resistance phenotype determination

*E. coli* isolates were collected from eight wastewater treatment utilities in Oregon as described previously (Khorshidi-Zadeh *et al*. submitted). Briefly, wastewater influent, secondary effluent, final effluent, and treated biosolids were collected from 17 wastewater treatment utilities over winter and summer in 2019 and 2020. To isolate *E. coli* colonies, collected samples were vacuum filtered or streaked directly onto m-TEC ChromoSelect agar (Sigma Aldrich, St. Louis, MO) plates and then confirmed by fluorescence on MacConkey agar with MUG (Hardy Diagnostics, Santa Maria, CA). Over the course of the study, 1143 *E. coli* colonies were isolated. Collected *E. coli* colonies were tested for the production of ESBL enzymes by measuring the zones of inhibition surrounding disks containing cefotaxime (30 μg), ceftazidime (30 μg), cefotaxime/clavulanic acid (30/10 μg), and ceftazidime/clavulanic acid (30/10 μg) (BD Diagnostics, Sparks, MD) (CLSI 2020). A difference in zone of inhibition of ≥ 5 mm diameter between antibiotic and antibiotic-acid disks indicated the production of ESBL enzymes. An internal quality control and *E. coli* ATCC 25922 strains were used as positive and negative controls, respectively. The phenotypic antibiotic resistance of the isolates to a series of beta-lactam antibiotics, including penicillins, 1^st^ – 4^th^ generation cephalosporins, and carbapenems were tested using the AST-GN99 card with a VITEK 2 system (bioMérieux, Marcy-l’Étoile, France) according to manufacturer’s instructions. The AST-GN99 card includes other classes of antibiotics (i.e., aminoglycosides, quinolones, tetracycline, nitrofuran, and sulfonamide) that supported the classification of multi-drug resistance phenotypes. Overall, 13 *E. coli* wastewater isolates were originally classified as ESBL-producers (Khorshidi-Zadeh *et al*. submitted); however, two of these isolates could not be confirmed as ESBL-producers using the AST-GN99 cards. Therefore, a total of 11 isolates were included in the present study. Details about the samples associated with these isolates are listed in Table S1.

### Whole genome sequencing and bioinformatic analysis

Freezer stocks were streaked onto tryptic soy agar (TSA; Hardy Diagnostics, Santa Maria, CA) and grown for 24 hours at 37 °C before being transferred to tryptic soy broth (TSB, Hardy Diagnostics, Santa Maria, CA). DNA was extracted from TSB cultures following manufacturer’s instructions for the DNeasy Blood and Tissue kit (Qiagen, Carlsbad, CA). Purified DNA was quantified and quality checked using a Qubit 4 (Invitrogen, Carslbad, CA) and Nanodrop (Thermo Fisher, Waltham, MA). DNA samples were submitted to the Oregon State University’s Center for Quantitative Life Sciences (Corvallis, OR) for library preparation using the plexWell™ 96 kit (seqWell, Beverly, MA). Libraries were sequenced on a MiSeq 3000 instrument (Illumina, San Diego, CA), employing paired-end 150 base reads. Duplicate libraries of each isolate were prepared to meet the minimum sample requirement of the plexWell™ 96 kit.

Demultiplexed reads were quality assessed using FastQC v0.11.8 (Andrews *et al*. 2012) and sequencing adapters were removed using Trimmomatic v0.40 (Bolger *et al*. 2014). Duplicate reads were concatenated into single forward and reverse FASTQ files and uploaded to the PATRIC platform for whole genome assembly and annotation using the “Comprehensive Genome Analysis” service (Davis *et al*. 2019). The resulting draft genome assemblies and annotation files were used for all downstream analyses.

### Multi-locus sequence typing and phylogenetic analysis

STs were determined from draft genome assemblies based on internal fragments of seven core genes (i.e., *adk, fumC, gyrB, icd, mdh, purA*, and *recA*) using the Center for Genomic Epidemiology (CGE) server (Larsen *et al*. 2012). A phylogenetic tree was created based on the concatenated sequences of the seven ST core genes’ fragments using the PhyML 3.1 Maximum-Likelihood model with 100 rounds of bootstrapping using the SeaView platform (Wirth *et al*. 2006; Gouy *et al*. 2010; Guindon *et al*. 2010).

### Identification of ARGs and virulence factors

ARGs were identified using NCBI’s AMRFinderPlus with minimum nucleotide identity of 90% and a minimum coverage of 50% (Feldgarden *et al*. 2019). Virulence factors were found using VirulenceFinder 2.0, with minimum nucleotide identity set to 90% and a minimum coverage of 60% for each (Joensen et al. 2014). Percent identities of amino acid sequences of AmpC were compared using Clustal Omega (Madeira *et al*. 2019).

### Data availability

The sequenced genomes have been deposited at the NCBI Sequence Read Archive (NCBI SRA) with the study identifier PRJNA767748.

## RESULTS

### Antibiotic resistance phenotypes of *E. coli* isolates

All 11 ESBL-producing *E. coli* isolates were resistant to at least one penicillin and several of the cephalosporin class beta-lactam antibiotics tested (Figure 1). In the penicillin class, all 11 *E. coli* isolates were resistant to ampicillin, and one isolate (G) was resistant to amoxicillin complexed with the beta-lactamase inhibitor clavulanic acid. Isolate G also demonstrated intermediate resistance to piperacillin/tazobactam complex, while all other isolates (*n* = 10) were susceptible. Six of the 11 *E. coli* isolates were resistant to all four generations of tested cephalosporins (i.e., first: cefazolin, second: cefuroxime, third: ceftriaxone, fourth: cefepime). The other five *E. coli* isolates carried phenotypic resistance to three generations of cephalosporins, while two demonstrated intermediate resistance to the fourth generation cefepime. None of the *E. coli* isolates displayed resistance to the tested carbapenems (i.e., ertapenem, imipenem, meropenem).

**Figure 1.**
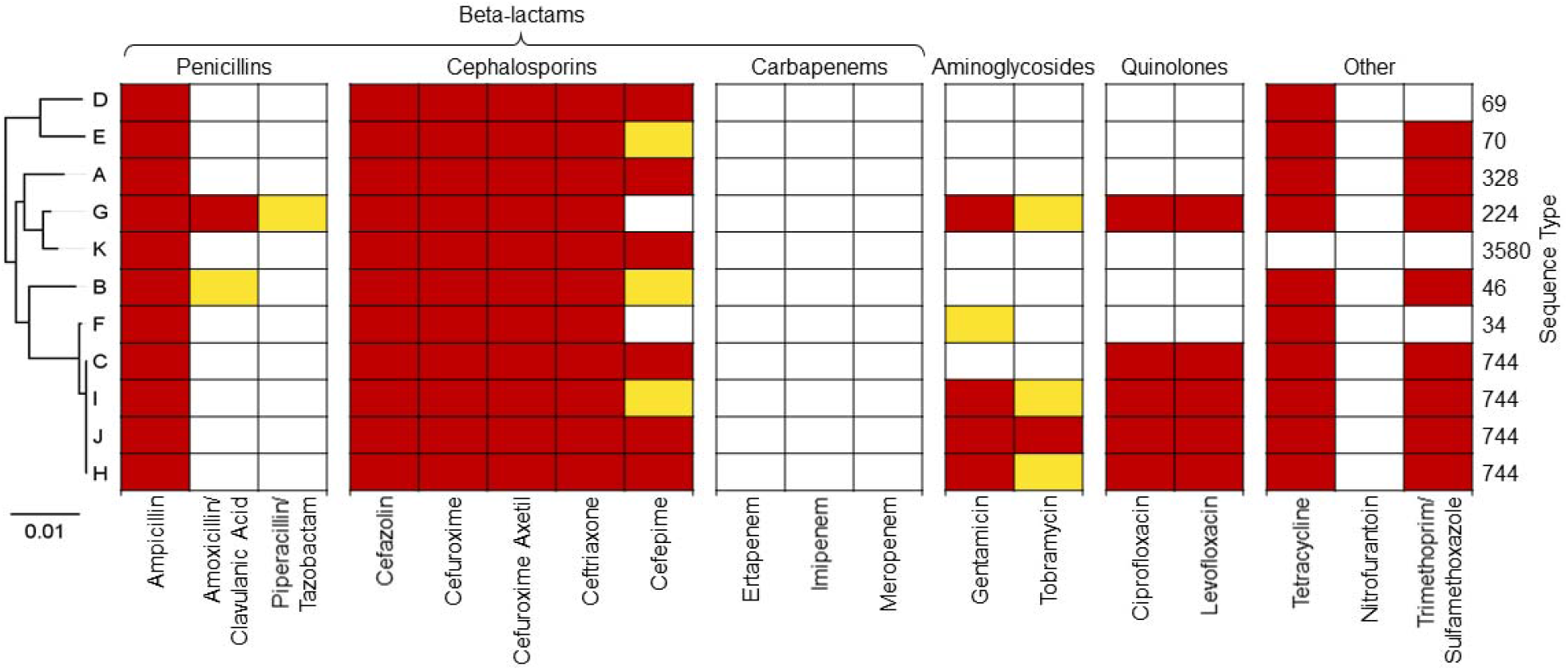
Antibiotic susceptibility phenotypes of ESBL-producing *E. coli* isolates to beta-lactams (penicillins, cephalosporins, and cabapenems) and to other classes of antibiotics. Antibiotic resistance classifications are shown in the table by dark red (resistant), yellow (intermediate), and white (susceptible). Sequence types (STs; right) are based on internal fragments of seven core genes (i.e., *adk, fumC, gyrB, icd, mdh, purA*, and *recA*). Phylogenetic tree (left) is based on concatenated sequences of the seven ST core genes’ fragments. Tree scale is in units of nucleotide substitutions per site.

Concerningly, eight of the 11 *E. coli* isolates were phenotypically multi-drug resistance (MDR) with resistances to three or more classes of antibiotics (Figure 1). Including beta-lactams, four of the *E. coli* isolates were phenotypically resistant to five classes of antibiotics (isolates G, I, J, and H). Tetracycline resistance was the most common detected non-beta-lactam antibiotic resistance (*n* = 10) and was observed in all the MDR *E. coli* isolates. Eight of the isolates were resistant to trimethoprim/sulfamethoxazole, but sensitive to nitrofurantoin. Five were resistant to both quinolones (ciprofloxacin and levofloxacin), and four were resistant to both aminoglycosides (gentamycin or tobramycin) tested.

### Genomic features of *E. coli* isolates

Genome sizes of the ESBL-producing isolates assemblies ranged from 4.59 Mbp (B) to 5.35 Mbp (A) with GC content between 50.3% (A) and 50.8% (F and G). ST analyses identified four of the 11 isolates to be ST744 (isolates C, H, I, and J; Figure 1). The seven other isolates were identified as singleton STs, including ST34, ST46, ST69, ST70, ST224, ST328, and ST3580 (Figure 1).

A total of 46 different virulence factors were identified in the sequenced data amongst the 11 *E. coli* isolates (Table S2). All 11 *E. coli* isolates contained the *terC* virulence factor, responsible for tellurite resistance (Valková *et al*. 2007). All *E. coli* isolates also carried the *gad* virulence factor, a glutamate decarboxylase that increases survival in acidic regions of the gastrointestinal tract (Damiano *et al*. 2015). The other common virulence factors were *sitA* (with a role in transportation of Fe^2+^ and Mn^2+^, *n* = 8) (Sabri *et al*. 2006), *ompT* (an outer membrane protein enabling intracellular survival, *n* = 6) (Hejair *et al*. 2017), and *traT* (transfers protein which inhibits certain pathways, *n* = 6) (Miajlovic and Smith 2014).

### ESBL-associated genes in the *E. coli* isolates

At least one known ESBL-associated gene (i.e., *bla*_CTX_, *bla*_TEM_, *bla*_CMY_, and *bla*_SHV_) was found in 10 of the 11 ESBL-producing *E. coli* isolates (Figures 2 and 3), and five of the isolates (A, B, G, I, and J) contained two of these genes. Eight *E. coli* isolates contained *bla*_CTX-M_ genes (Figure 2a). Four of the eight isolates with *bla*_CTX-M_ gene – isolates C, H, I, and J which were all identified as ST744. These four *E. coli* isolates harbored arsenic resistance genes nearby the *bla*_CTX-M_ gene (labeled 2 and 3 in Figure 2a). Six of the *bla*_CTX-M_ genes were identified as subtype *bla*_CTX-M-55_ (isolates B, C, D, H, I, and J) and two were identified as subtype *bla*_CTX-M-15_ (A and K). All six of the *bla*_CTX-M-55_ carrying isolates were flanked by the same mobile genetic elements (i.e., transposase, labeled 1 in Figure 2a). The notably different gene region was from isolate D, which was identified as subtype *bla*_CTX-M-55_ adjacent to a set of the toxin-antitoxin *yee* genes and near a death-on-curing *(doc*) gene (Lehnherr *et al*. 1993; Brown and Shaw 2003).

**Figure 2.**
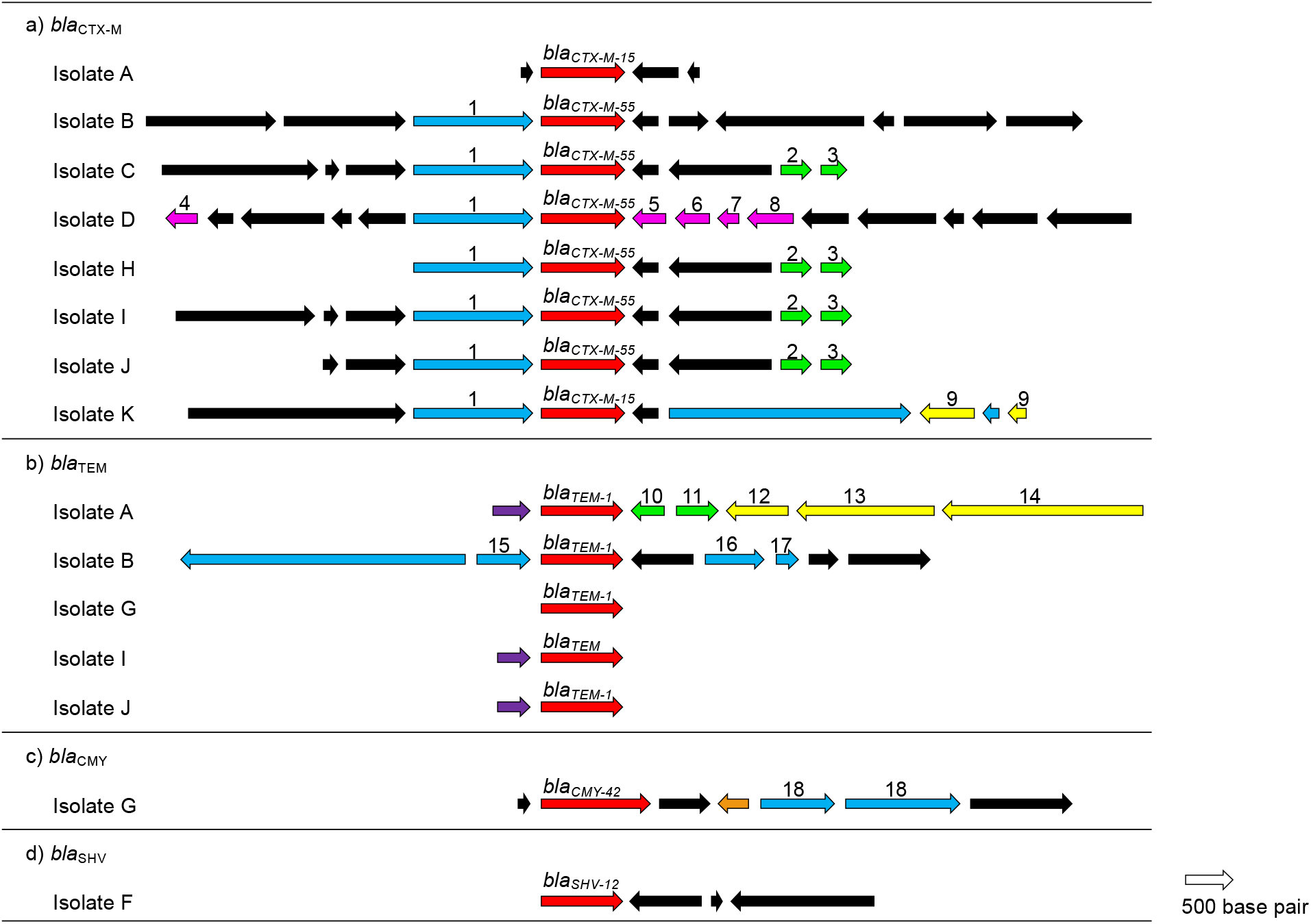
Gene regions for *bla*_CTX-M_, *bla*_TEM_, *bla*_CMY_, and *bla*_SHV_. Arrows indicate the direction of translation, and colors indicate gene functionality (red: ESBL-encoding genes, blue: mobile genetic elements, green: heavy metal associated, yellow: *Tn7* transposon associated, pink: possible plasmid addiction system related, purple: phage integrase associated, orange: multidrug resistance efflux pump, black: other functions). 1. transposes, 2. arsenical resistance operon, 3. arsenic resistance protein *arsH*, 4. *doc* toxin, 5. *YeeV* toxin protein, 6. *YeeU* protein (antitoxin to *YeeV*), 7. *YeeT* protein, 8. *YeeS* protein, 9. transposase encoding genes, 10. mercuric transport protein, *MerT*, 11. mercuric resistance operon, 12. *Tn7*-like transposition protein D, 13. *Tn7*-like transposition protein C, 14. *Tn7*-like transposition protein B, 15. transposon resolvase, 16. plasmid partitioning protein *ParA*, 17. plasmid partition protein *ParG*, 18. conjugal transfer protein encoding. Gene lengths are displayed to scale.

**Figure 3.**
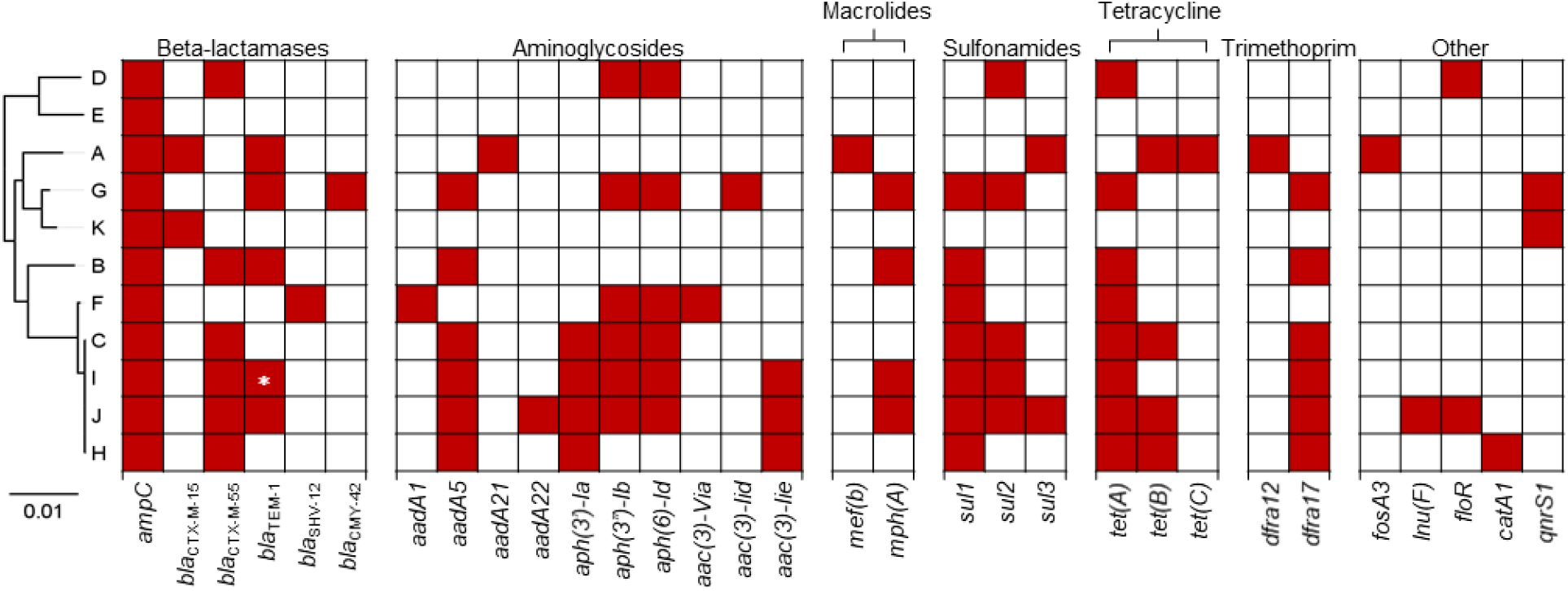
Antibiotic resistance genes identified in the genomes of ESBL-producing *E. coli* isolated from wastewater facilities in Oregon. Antibiotic resistance genes prevalence is shown in the table by dark red (present) and white (absent). Asterisk (*) highlights the identified gene as TEM.

*bla*_TEM_ genes were identified in five of the ESBL-producing *E. coli* isolates (Figure 2b and 3). Unlike the *bla*_CTX-M_ gene regions, the *bla*_TEM_ gene regions were not flanked by transposases. However, three of the five *bla*_TEM_-containing isolates identified *bla*_TEM_ adjacent to a phage integrase gene (purple arrows in Figure 2b). Moreover, *bla*_CMY_ gene was found within only one of the isolates (G), reported as a subtype *bla*_CMY-42_ (Figure 2c). In addition to the *bla*_CMY_ gene, isolate G also carried the *bla*_TEM-1_ gene. A *bla*_SHV_ gene was found in isolate F and was reported as subtype *bla*_SHV-12_ (Figure 2d). The *bla*_CMY_ and *bla*_SHV_ gene regions are similar to respective reference gene regions from other bacteria in the PATRIC database. Despite the multidrug resistance phenotype of many of the isolates, only isolate G had a multidrug resistance related gene (efflux pump) nearby the ESBL gene (orange arrow in Figure 2c).

### Other ARGs in the *E. coli* isolates

The analysis of sequenced data via AMRFinderPlus identified a total of 31 different ARGs belonging to 11 classes of antibiotics (Figure 3). All the 11 ESBL-producing *E. coli* isolates contained *ampC*. AmpC beta-lactamases are cephalosporinases of clinical importance as they confer resistance to cephalothin, cefazolin, cefoxitin, most penicillins, and beta-lactamase inhibitor-beta-lactam complexes (Jacoby 2009). Results show very conserved location for *ampC*. The average nucleotide identity matrix between the *E. coli* isolates for the *ampC* is shown in Table S3. Overall, *ampC* genes ranged from 96.30 to 100.0% nucleotide identity. Unsurprising, isolates C, H, I, and J that were all identified as ST744 showed 100.0% similarity for *ampC*. Moreover, nine of the 11 *E. coli* isolates contained genes encoding for aminoglycoside resistance, where *aadA5, aph(3’’)-Ib*, and *aph(6)-Id* were the most common genes (*n* = 6; Figure 3). Those same nine isolates with resistance to aminoglycosides also contained resistances to sulfonamides and tetracyclines. The most common sulfonamide resistance gene was *sul1 (n* = 7), followed by *sul2 (n* = 5). The most common tetracycline resistance gene was *tet(A)* (*n* = 8). Seven *E. coli* isolates harbored a trimethoprim resistance gene, six of which harbored *drfA17* and only one carried *drfA12*. Genes encoding resistances to fosfomycins, chloramphenicols, quinolones, and lincosamides were also detected among the *E. coli* isolates (Figure 3).

Comparing the resistance phenotypes and genotypes of the 11 ESBL-producing *E. coli* isolates, some similarities and discrepancies were observed. All isolates harbored phenotypic resistance to the tested first, second, and third generation cephalosporins (i.e., first: cefazolin, second: cefuroxime, third: ceftriaxone; Figure 1), while all but one (isolate E) carried at least one of the known ESBL genes (Figure 3). The lack of any ESBL-associated gene in isolate E suggests presence of another novel mechanism supporting cephalosporin resistance. Moreover, the two *E. coli* isolates G and F that remained susceptible to cefepime (fourth generation cephalosporin) did not carry the *bla*_CTX-M_ gene (Figure 3), which suggests a correlation between prevalence of *bla*_CTX_ gene and fourth generation cephalosporins. Moreover, all 11 *E. coli* isolates harbored the *ampC* gene (Figure 3) and AmpC beta-lactamases commonly mediate resistance to beta-lactamase inhibitor-beta-lactam combinations; however, only one isolate (G) showed resistance to amoxicillin/clavulanic acid, and one isolate (B) had intermediate resistance (Figure 1). Comparing resistances to aminoglycosides, results suggest the *aadA5* gene (Figure 3) as a good indicator for prevalence of phenotypic resistances to gentamicin and tobramycin (Figure 1). For resistance to tetracycline, we observed phenotypic resistance of all but one *E. coli* isolate (K) (Figure 1) and found one of three resistance genes (*tet(A), tet(B)*, and *tet(C)*) in all but isolates E and K, which shows these three tetracycline resistance genes as good indicators of their associated resistance phenotype.

## DISCUSSION

Global spread of pathogens resistant to antibiotic treatments is a major public health issue. According to the most recent CDC report on antibiotic resistance, over 2.8 million cases of infections in the U.S. are caused by antibiotic-resistant bacteria with over 35,000 per year associated deaths (CDC, 2019). Resistance via ESBL-producing bacteria is especially concerning because of the vast number of antibiotics they confer resistance to (i.e., penicillins and cephalosporins). It is estimated that healthcare complication risk is increased by 50% if infections are caused by ESBL-producing Enterobacteriaceae (CDC, 2019). Last-resort beta-lactam antibiotics like carbapenems are currently used for the treatment of ESBL-producing *E. coli* infections (Hawkey and Livermore 2012); however, certain ESBL-associated genes (e.g., *bla*_VIM_ and *bla*_KPC_) have been shown to mediate resistance to these last resort antibiotics (Hoelle *et al*. 2019).

Few studies have characterized ESBL determinant genes in *E. coli* isolated from wastewater treatment utilities in the U.S. One such study, limited to isolates resistant to the imipenem, found a variety of ESBL-related genes including *bla*_CTX-M_, *bla*_SHV_, and *bla*_TEM_ to be prevalent in wastewater samples from seven states (Hoelle *et al*. 2019). Another study reported that ESBL-producing *E. coli* isolated from Colorado wastewater treatment plants expressed distinct genetic profiles compared to sewage, with wastewater samples more likely to carry *bla*_CTX_ type genes and more likely to be MDR (Haberecht *et al*. 2019). Our results agree with those findings.

In this study, 11 ESBL-producing *E. coli* isolated from multiple wastewater treatment utilities throughout Oregon were characterized for their genetic determinants. Many of these isolates harbored MDR phenotypic resistances and carried determinant genes for a wide array of antibiotics in addition to beta-lactams including tetracycline, aminoglycosides, and quinolones. Wastewater treatment utilities are an excellent setting for the study of antibiotic resistance because they represent a composite sample of the community. The common MDR phenotypes in most of the isolates in this study is concerning. Moreover, four of 11 *E. coli* isolates (isolates C, H, I, and J) that were all identified as ST744 carried 100.0% similarity between their average nucleotide identities for *ampC* while also having very similar synteny for the *bla*_CTX-M-55_ gene.

Moreover, findings showed one isolate (E) with an ESBL-associated phenotype (resistance to first, second, and third generation cephalosporins) without harboring any of the known ESBL genes. This discrepancy shows the potential limitation of annotation tools for uncharacterized novel genes that confer beta-lactam resistance. As shown by the isolates encompassing multiple STs, the ESBL-positive *E. coli* isolated from Oregon wastewater treatment plants were phylogenetically diverse. However, the same few ESBL genes were found across multiple STs with high similarities between their average nucleotide identities of *ampC*. The dissociation of core genome diversity and resistance genotypes is indicative of horizontal gene transfer. This is also supported by the colocalization of mobile genetic elements with the ESBL genes, especially *bla*_CTX-M_ types. The spread of beta-lactam resistance is likely not limited to conjugation via a plasmid as determinant genes could be transposed to other plasmids or chromosomes. The co-occurrences of the ESBL-associated genes with other ARGs as well as several virulence factors in these ESBL-producing *E. coli* isolates is concerning. Given that these isolates were collected from multiple wastewater treatment utilities in Oregon, these results indicate that ESBL resistance occurs in Oregon’s wastewater systems, and the associated resistance is potentially transferable to other reservoirs, including receiving rivers and agricultural zones utilizing biosolids. Results confirm the need for further characterization of ESBL-producing Enterobacteriaceae resistome, especially in wastewater systems where confluence of municipal and clinical waste streams provide an ideal environment for the spread of ARGs.

## CONCLUSIONS

This study characterizes the genotypes and phenotypes of antibiotic resistance in 11 ESBL-producing *E. coli* isolates collected from Oregon wastewater systems. Known ESBL-associated genes (i.e., *bla*_*CMY*_, *bla*_*CTX*_, *bla*_*SHV*_, *bla*_*TEM*_) were identified in 10 of the 11 *E. coli* isolates with five isolates carrying two of these genes. The *ampC* gene and virulence factors were observed in all the *E. coli* isolates. Moreover, 31 different ARGs were identified across all the isolates. All *E. coli* isolates harbored phenotypic resistance to beta-lactams with eight isolates demonstrating MDR phenotypes. Six of the eleven isolates were sourced from wastewater products (biosolids and final effluents). These results demonstrate the risks of ESBL-producing *E. coli* in wastewater systems that can enter the environment posing a critical threat to public health.

## Data Availability

All data produced are available online at the NCBI Sequence Read Archive (NCBI SRA) with the study identifier PRJNA767748.

## REFERENCES

Alexander, J., Hembach, N. & Schwartz, T., 2020. Evaluation of antibiotic resistance dissemination by wastewater treatment plant effluents with different catchment areas in Germany. Scientific Reports, 10.

Andrews, S., Krueger, F., Segonds-Pichon, A., Biggins, L., Kruger, C. & Wingett, S., 2012. FastQC. Babraham Bioinformatics.

Ben Said, L., Jouini, A., Klibi, N., Dziri, R., Alonso, C. A., Boudabous, A., Ben Slama, K. & Torres, C., 2015. Detection of extended-spectrum beta-lactamase (ESBL)-producing Enterobacteriaceae in vegetables, soil and water of the farm environment in Tunisia. International Journal of Food Microbiology, 203, 86–92.

Blaak, H., Lynch, G., Italiaander, R., Hamidjaja, R. A., Schets, F. M. & de Roda Husman, A. M., 2015. Multidrug-Resistant and Extended Spectrum Beta-Lactamase-Producing Escherichia coli in Dutch Surface Water and Wastewater. (Mokrousov, I., Ed.)PLOS ONE, 10, e0127752.

Bolger, A. M., Lohse, M. & Usadel, B., 2014. Trimmomatic: a flexible trimmer for Illumina sequence data. Bioinformatics, 30, 2114–2120.

Bradford, P. A., 2001. Extended-Spectrum □-Lactamases in the 21st Century: Characterization, Epidemiology, and Detection of This Important Resistance Threat. CLIN. MICROBIOL. REV., 14, 19.

Brown, J. M. & Shaw, K. J., 2003. A Novel Family of Escherichia coli Toxin-Antitoxin Gene Pairs. Journal of Bacteriology, 185, 6600–6608.

Bush, K. & Fisher, J. F., 2011. Epidemiological Expansion, Structural Studies, and Clinical Challenges of New β-Lactamases from Gram-Negative Bacteria. Annual Review of Microbiology, 65, 455–478.

Centers for Disease Control and Prevention (U.S.), 2019. Antibiotic resistance threats in the United States, 2019. Centers for Disease Control and Prevention (U.S.).

CLSI, 2020. M100 Performance Standards for Antimicrobial Susceptibility Testing A CLSI supplement for global application. 30th Edition.

Damiano, M. A., Bastianelli, D., Al Dahouk, S., Köhler, S., Cloeckaert, A., De Biase, D. & Occhialini, A., 2015. Glutamate Decarboxylase-Dependent Acid Resistance in Brucella spp.: Distribution and Contribution to Fitness under Extremely Acidic Conditions. Applied and Environmental Microbiology, 81, 578–586.

Davis, J. J., Wattam, A. R., Aziz, R. K., Brettin, T., Butler, R., Butler, R. M., Chlenski, P., Conrad, N., Dickerman, A., Dietrich, E. M., Gabbard, J. L., Gerdes, S., Guard, A., Kenyon, R. W., Machi, D., Mao, C., Murphy-Olson, D., Nguyen, M., Nordberg, E. K. et al., 2019. The PATRIC Bioinformatics Resource Center: expanding data and analysis capabilities. Nucleic Acids Research, gkz943.

Feldgarden, M., Brover, V., Haft, D. H., Prasad, A. B., Slotta, D. J., Tolstoy, I., Tyson, G. H., Zhao, S., Hsu, C. H., McDermott, P. F., Tadesse, D. A., Morales, C., Simmons, M., Tillman, G., Wasilenko, J., Folster, J. P. & Klimke, W., 2019. Validating the AMRFinder Tool and Resistance Gene Database by Using Antimicrobial Resistance Genotype-Phenotype Correlations in a Collection of Isolates. Antimicrobial Agents and Chemotherapy, 63.

Gouy, M., Guindon, S. & Gascuel, O., 2010. SeaView Version 4: A Multiplatform Graphical User Interface for Sequence Alignment and Phylogenetic Tree Building. Molecular Biology and Evolution, 27, 221–224.

Guindon, S., Dufayard, J. F., Lefort, V., Anisimova, M., Hordijk, W. & Gascuel, O., 2010. New Algorithms and Methods to Estimate Maximum-Likelihood Phylogenies: Assessing the Performance of PhyML 3.0. Systematic Biology, 59, 307–321.

Haberecht, H. B., Nealon, N. J., Gilliland, J. R., Holder, A. V., Runyan, C., Oppel, R. C., Ibrahim, H. M., Mueller, L., Schrupp, F., Vilchez, S., Antony, L., Scaria, J. & Ryan, E. P., 2019. Antimicrobial-Resistant Escherichia coli from Environmental Waters in Northern Colorado. Journal of Environmental and Public Health, 2019, 1–13.

Haenni, M., Beyrouthy, R., Lupo, A., Châtre, P., Madec, J. Y. & Bonnet, R., 2018. Epidemic spread of Escherichia coli ST744 isolates carrying mcr-3 and blaCTX-M-55 in cattle in France. Journal of Antimicrobial Chemotherapy, 73, 533–536.

Haghighatpanah, M., Mozaffari Nejad, A. S., Mojtahedi, A., Amirmozafari, N. & Zeighami, H., 2016. Detection of extended-spectrum β-lactamase (ESBL) and plasmid-borne blaCTX-M and blaTEM genes among clinical strains of Escherichia coli isolated from patients in the north of Iran. Journal of Global Antimicrobial Resistance, 7, 110–113.

Hawkey, P. M. & Livermore, D. M., 2012. Carbapenem antibiotics for serious infections. BMJ, 344, e3236–e3236.

Hejair, H. M. A., Ma, J., Zhu, Y., Sun, M., Dong, W., Zhang, Y., Pan, Z., Zhang, W. & Yao, H., 2017. Role of outer membrane protein T in pathogenicity of avian pathogenic Escherichia coli. Research in Veterinary Science, 115, 109–116.

Hoelle, J., Johnson, J. R., Johnston, B. D., Kinkle, B., Boczek, L., Ryu, H. & Hayes, S., 2019. Survey of US wastewater for carbapenem-resistant Enterobacteriaceae. Journal of Water and Health, 17, 219–226.

Jacoby, G. A., 2009. AmpC β-Lactamases. Clinical Microbiology Reviews, 22, 161–182.

Jiang, X., Cui, X., Xu, H., Liu, W., Tao, F., Shao, T., Pan, X. & Zheng, B., 2019. Whole genome sequencing of extended-spectrum beta-lactamase (ESBL)-producing escherichia coli isolated from a wastewater treatment plant in China. Frontiers in Microbiology, 10.

Joensen, K. G., Scheutz, F., Lund, O., Hasman, H., Kaas, R. S., Nielsen, E. M. & Aarestrup, F. M., 2014. Real-Time Whole-Genome Sequencing for Routine Typing, Surveillance, and Outbreak Detection of Verotoxigenic Escherichia coli. Journal of Clinical Microbiology, 52, 1501–1510.

Jørgensen, S. B., Søraas, A. V., Arnesen, L. S., Leegaard, T. M., Sundsfjord, A. & Jenum, P. A., 2017. A comparison of extended spectrum β-lactamase producing Escherichia coli from clinical, recreational water and wastewater samples associated in time and location. (Singer, A. C., Ed.) PLOS ONE, 12, e0186576.

Khorshidi-Zadeh, M., Yiu, S. Y., Nguyen, J. N., Garza, G. L., Waite-Cusic, J., Radniecki, T. R. & Navab-Daneshmand, T., submitted. Antibiotic resistance profile of E. coli isolates in 17 municipal wastewater utilities across Oregon.

Larsen, M. V., Cosentino, S., Rasmussen, S., Friis, C., Hasman, H., Marvig, R. L., Jelsbak, L., Sicheritz-Pontén, T., Ussery, D. W., Aarestrup, F. M. & Lund, O., 2012. Multilocus sequence typing of total-genome-sequenced bacteria. Journal of Clinical Microbiology, 50, 1355–1361.

Lehnherr, H., Maguin, E., Jafri, S. & Yarmolinsky, M. B., 1993. Plasmid Addiction Genes of Bacteriophage P1: doc, which Causes Cell Death on Curing of Prophage, and phd, which Prevents Host Death when Prophage is Retained. Journal of Molecular Biology, 233, 414–428.

Liu, G., Bogaj, K., Bortolaia, V., Olsen, J. E. & Thomsen, L. E., 2019. Antibiotic-Induced, Increased Conjugative Transfer Is Common to Diverse Naturally Occurring ESBL Plasmids in Escherichia coli. Frontiers in Microbiology, 10.

Madeira, F., Park, Y. mi, Lee, J., Buso, N., Gur, T., Madhusoodanan, N., Basutkar, P., Tivey, A. R. N., Potter, S. C., Finn, R. D. & Lopez, R., 2019. The EMBL-EBI search and sequence analysis tools APIs in 2019. Nucleic Acids Research, 47, W636–W641.

Manaia, C. M., Rocha, J., Scaccia, N., Marano, R., Radu, E., Biancullo, F., Cerqueira, F., Fortunato, G., Iakovides, I. C., Zammit, I., Kampouris, I., Vaz-Moreira, I. & Nunes, O. C., 2018. Antibiotic resistance in wastewater treatment plants: Tackling the black box. Environment International, 115, 312–324.

Miajlovic, H. & Smith, S. G., 2014. Bacterial self-defence: how Escherichia coli evades serum killing. FEMS Microbiology Letters, 354, 1–9.

Palucha, A., Mikiewicz, B., Hryniewicz, W. & Gniadkowski, M., 1999. Concurrent outbreaks of extended-spectrum β-lactamase-producing organisms of the family Enterobacteriaceae in a Warsaw hospital. Journal of Antimicrobial Chemotherapy, 44, 489–499.

Park, Y., Kang, H. K., Bae, I. K., Kim, J., Kim, J. S., Uh, Y., Jeong, S. H. & Lee, K., 2009. Prevalence of the Extended-Spectrum β -Lactamase and qnr Genes in Clinical Isolates of Escherichia coli. Annals of Laboratory Medicine, 29, 218–223.

Paulshus, E., Thorell, K., Guzman-Otazo, J., Joffre, E., Colque, P., Kühn, I., Möllby, R., Sørum, H. & Sjöling, Å., 2019. Repeated Isolation of Extended-Spectrum-β-Lactamase-Positive Escherichia coli Sequence Types 648 and 131 from Community Wastewater Indicates that Sewage Systems Are Important Sources of Emerging Clones of Antibiotic-Resistant Bacteria.

Riquelme, M. V., Garner, E., Gupta, S., Metch, J., Zhu, N., Blair, M. F., Arango-Argoty, G., Maile-Moskowitz, A., Li, A., Flach, C. F., Aga, D. S., Nambi, I., Larsson, D. G. J., Bürgmann, H., Zhang, T., Pruden, A. & Vikesland, P. J., 2021. Wastewater Based Epidemiology Enabled Surveillance of Antibiotic Resistance, p. 2021.06.01.21258164.

Sabri, M., Léveillé, S. & Dozois, C. M. Y., 2006. A SitABCD homologue from an avian pathogenic Escherichia coli strain mediates transport of iron and manganese and resistance to hydrogen peroxide. Microbiology, 152, 745–758.

Schages, L., Wichern, F., Kalscheuer, R. & Bockmühl, D., 2020. Winter is coming – Impact of temperature on the variation of beta-lactamase and mcr genes in a wastewater treatment plant. Science of The Total Environment, 712, 136499.

Schmiege, D., Zacharias, N., Sib, E., Falkenberg, T., Moebus, S., Evers, M. & Kistemann, T., 2021. Prevalence of multidrug-resistant and extended-spectrum beta-lactamase-producing Escherichia coli in urban community wastewater. Science of The Total Environment, 785, 147269.

Schwaber, M. J., Navon-Venezia, S., Schwartz, D. & Carmeli, Y., 2005. High levels of antimicrobial coresistance among extended-spectrum-β-lactamase-producing Enterobacteriaceae. Antimicrobial Agents and Chemotherapy, 49, 2137–2139.

Schwaber, M. J., Navon-Venezia, S., Kaye, K. S., Ben-Ami, R., Schwartz, D. & Carmeli, Y., 2006. Clinical and Economic Impact of Bacteremia with Extended-Spectrum-β-Lactamase-Producing Enterobacteriaceae. Antimicrobial Agents and Chemotherapy, 50, 1257–1262.

Severin, J. A., Mertaniasih, N. M., Kuntaman, K., Lestari, E. S., Purwanta, M., Lemmens-Den Toom, N., Duerink, D. O., Hadi, U., van Belkum, A., Verbrugh, H. A. & Goessens, W. H., 2010. Molecular characterization of extended-spectrum β-lactamases in clinical Escherichia coli and Klebsiella pneumoniae isolates from Surabaya, Indonesia. Journal of Antimicrobial Chemotherapy, 65, 465–469.

Sidjabat, H. E., Paterson, D. L., Adams-Haduch, J. M., Ewan, L., Pasculle, A. W., Muto, C. A., Tian, G. B. & Doi, Y., 2009. Molecular Epidemiology of CTX-M-Producing Escherichia coli Isolates at a Tertiary Medical Center in Western Pennsylvania. Antimicrobial Agents and Chemotherapy, 53, 4733–4739.

Turkovicova, L., Smidak, R., Jung, G., Turna, J., Lubec, G. & Aradska, J., 2016. Proteomic analysis of the TerC interactome: Novel links to tellurite resistance and pathogenicity. Journal of Proteomics, 136, 167–173.

Valková, D., Valkovičová, L., Vávrová, S., Kováčová, E., Mravec, J. & Turňa, J., 2007. The contribution of tellurite resistance genes to the fitness of Escherichia coli uropathogenic strains. Open Life Sciences, 2, 182–191.

Wirth, T., Falush, D., Lan, R., Colles, F., Mensa, P., Wieler, L. H., Karch, H., Reeves, P. R., Maiden, M. C., Ochman, H. & Achtman, M., 2006. Sex and virulence in Escherichia coli: an evolutionary perspective. Molecular Microbiology, 60, 1136–1151.

