## Supplementary material for "Resistome characterization of extended-spectrum beta-lactamase (ESBL)-producing *Escherichia coli* isolated from wastewater treatment utilities in Oregon"

Short title: Characterizing resistome of ESBL-producing *E. coli* from Oregon wastewater

Maeghan Easler^a,†^, Clinton Cheney^a,†^, Jared D. Johnson^b^, Marjan Khorshidi Zadeh^a^, Jacquelynn N. Nguyen^a^, Sue Yee Yiu^a^, Joy Waite-Cusic^b^, Tyler S. Radniecki^a^, Tala Navab-Daneshmand^a,*^

^a^ 105 SW 26^th^ St, 116 Johnson Hall, School of Chemical, Biological, and Environmental Engineering, Oregon State University, Corvallis, OR 97331, United States

^b^ 3051 SW Campus Way, Department of Food Science and Technology, Oregon State University, Corvallis, OR 97331, United States

^†^ These authors contributed equally to this work.

Authors’ email addresses:

Maeghan Easler:, Clinton Cheney:, Jared D. Johnson:, Marjan Khorshidi Zadeh:, Jacquelynn N. Nguyen:, Sue Yee Yiu:, Joy Waite-Cusic:, Tyler S. Radniecki:, Tala Navab-Daneshmand:

**Supplementary Table 1.** Source characterization of ESBL-producing *E. coli* isolates collected from wastewater treatment utilities across the state of Oregon.

| *E. coli* isolate ID | Utility ID | Collection season | Wastewater flow |
| --- | --- | --- | --- |
| A | 1 | Winter 2020 | Influent |
| B | 1 | Summer 2020 | Influent |
| C | 2 | Winter 2019 | Biosolids |
| D | 3 | Summer 2020 | Biosolids |
| E | 3 | Winter 2019 | Secondary Effluent |
| F | 4 | Summer 2019 | Final effluent |
| G | 5 | Winter 2020 | Secondary Effluent |
| H | 6 | Summer 2020 | Biosolids |
| I | 7 | Winter 2020 | Final effluent |
| J | 7 | Summer 2020 | Influent |
| K | 8 | Winter 2020 | Final effluent |

**Supplementary Table 2.** Virulence factors identified in ESBL-producing *E. coli* isolated from wastewater flows (i.e., influent, secondary effluent, final effluent, and biosolids) in Oregon.

| *E. coli* isolates | Virulence factors |
| --- | --- |
| A | *cif, eae, efa1, espA, espB, espF, espJ, gad, iss, lpfA, nleA, nleB, nleC, ompT, terC, tir, traT* |
| B | *astA, cma, cvaC, gad, hlyF, iucC, iutA, ompT, sitA, terC, traT* |
| C | *air, chuA, eilA, gad, hra, iss, kpsE, kpsMII_K5, lpfA, papA_F19, papC, terC* |
| D | *air, chuA, cib, eilA, fyuA, gad, irp2, kpsE, ompT, sitA, terC* |
| E | *gad, iss, sitA, terC* |
| F | *cib, cma, cvaC, gad, hlyF, iroN, iss, ompT, sitA, terC, traT* |
| G | *gad, lpfA, terC* |
| H | *afaA, afaB, afaC, afaD, afaE8, gad, iucC, iutA, sitA, terC, traT* |
| I | *cma, cvaC, gad, hlyF, iucC, iutA, ompT sitA, terC, traT* |
| J | *afaA, afaB, afaC, afaD, afaE8, cma, gad, hlyF, hra, ireA, iucC, iutA, mcmA, ompT, papA_F11, papC, sitA, terC, traT* |
| K | *cba, cia, cma, gad, lpfA, sitA, terC* |

**Supplementary Table 3.** Average nucleotide identity between 11 ESBL-producing *E. coli* isolated from wastewater flows (i.e., influent, secondary effluent, final effluent, and biosolids) in Oregon.

|  | A |  |  |  |  |  |  |  |  |  |  |
| --- | --- | --- | --- | --- | --- | --- | --- | --- | --- | --- | --- |
| A | 100.00 | B |  |  |  |  |  |  |  |  |  |
| B | 99.29 | 100.00 | C |  |  |  |  |  |  |  |  |
| C | 97.71 | 97.35 | 100.00 | D |  |  |  |  |  |  |  |
| D | 96.56 | 96.56 | 97.97 | 100.00 | E |  |  |  |  |  |  |
| E | 96.91 | 97.09 | 98.15 | 98.24 | 100.00 | F |  |  |  |  |  |
| F | 99.21 | 99.91 | 97.44 | 96.65 | 97.18 | 100.00 | G |  |  |  |  |
| G | 99.82 | 99.12 | 97.53 | 96.38 | 96.74 | 99.03 | 100.00 | H |  |  |  |
| H | 97.71 | 97.35 | 100.00 | 97.97 | 98.15 | 97.44 | 97.53 | 100.00 | I |  |  |
| I | 97.71 | 97.35 | 100.00 | 97.97 | 98.15 | 97.44 | 97.53 | 100.00 | 100.00 | J |  |
| J | 97.71 | 97.35 | 100.00 | 97.97 | 98.15 | 97.44 | 97.53 | 100.00 | 100.00 | 100.00 | K |
| K | 99.12 | 98.94 | 97.18 | 96.30 | 97.18 | 99.03 | 99.12 | 97.18 | 97.18 | 97.18 | 100.00 |
